# Probiotics Reduce Group B Streptococcus (GBS) Colonization During Pregnancy: A Systematic Review and Meta-Analysis

**DOI:** 10.64898/2026.05.01.26352246

**Authors:** Amber Broughton, Kevin Rowland, David Raskin

## Abstract

**Background:** Transfer of *Streptococcus agalactiae*, or Group B Streptococcus (GBS) from parent to newborn during delivery can produce life-threatening infections in neonates. Probiotics could potentially prevent GBS colonization in pregnant individuals. We conducted a systematic review and meta-analysis to evaluate the effectiveness of probiotic administration in treating Group B Streptococcus colonization.

**Methods:** MEDLINE, ClinicalTrials.gov, PROSPERO, and the Cochrane, Wild Card, Central Register of Controlled Trials were searched from the July 2015 of each database until July 2025 that completed a randomized controlled trial which compared Probiotic versus control. We utilized the Cochrane Risk of Bias 2.0 (RoB 2) tool to assess bias in the systematic review.

**Results:** 14 randomized controlled clinical trials met our inclusion criteria. The trials used oral probiotic administration compared to either a placebo or a control group. A meta-analysis showed that probiotic administration produced a statistically significant decrease in the rate of GBS colonization in pregnant individuals. The individual studies ranged from four showing great effectiveness, while the other 10 studies showed a range of effectiveness, from partially effective to no effectiveness in preventing GBS colonization.

**Conclusion:** Overall, probiotics were effective in lowering infection rates of GBS, but individual studies showed great variability. Probiotics show promise in decreasing GBS colonization in pregnant people, but more studies need to be performed in order to use them effectively and decrease antibiotic usage.

## INTRODUCTION

*Streptococcus agalactiae*, also known as Group B Streptococcus (GBS), is a Gram-positive beta-hemolytic bacterium. that is sometimes found in the vaginal microbiome. GBS commonly colonizes the genitourinary tract and gastrointestinal tract [1]. In healthy adults, GBS is usually harmless, but in newborns, it can produce life-threatening infections, including septicemia, meningitis, and endocarditis [2,3]. Secondary adverse events include neonatal prematurity, developmental delay, and stillbirth [3]. Transmission to newborns occurs during labor, due to ruptured membranes in the childbirth process [3].

The American College of Obstetricians and Gynecologists (ACOG) recommends that pregnant individuals be tested for GBS colonization through rectovaginal swab at 35-38 weeks of pregnancy [1]. According to the United States Centers for Disease Control and Prevention, 1 in 4 pregnant individuals will test positive for GBS. Current clinical guidelines recommend intrapartum intravenous penicillin as the first-line prophylaxis for all pregnant individuals who test positive for Group B *Streptococcus* (GBS) colonization. Utilizing antibiotics for prophylaxis can produce adverse effects, including diarrhea, *Clostridioides difficile* infections in patients, and anaphylaxis [4]. In addition, using antibiotic prophylaxis to prevent neonatal meningitis may lead to new antibiotic resistance in meningitis-causing bacteria [5].

Probiotic treatments have the potential to protect against GBS infections without antibiotic-related side effects. Many recent clinical trials have tested whether probiotics are effective in decreasing the rate of GBS colonization in pregnant individuals. The objective of this systematic review and meta-analysis is to understand the efficacy of probiotics in preventing colonization of GBS by analyzing the data of recently published randomized clinical trials. We then performed a meta-analysis using all studies that fit our criteria to analyze effectiveness of probiotics in preventing GBS colonization factoring in all studies.

## METHODS

### Inclusion Criteria

Selection criteria included all Randomized Clinical Trials that investigated the effects of probiotics on GBS colonization. We included studies that were focused on pregnant people receiving probiotics (any strain, dose, route, duration) aimed at reducing GBS colonization. The intervention had to be probiotics (defined as live microorganisms that have beneficial effects on the body), utilizing any probiotic strain(s), dosage, or route of administration. The studies had to include a comparator group which could be a placebo, no treatment, or standard care. The studies were included if they reported on rectovaginal Group B Streptococcus colonization status, which could be either positive or negative. Studies could also report on secondary outcome measures including NICU admission and neonatal infection, maternal chorioamnionitis, preterm labor, preterm birth (< 37 weeks gestational age) adverse reactions for intrapartum antibiotics.

### Exclusion Criteria

Studies other than Randomized Controlled Trials were excluded. In vitro and animal studies were also excluded. Studies that did not involve pregnant people or if antibiotics were administered for reasons other than GBS colonization were also excluded. Studies that did not report on GBS colonization or our secondary outcome measures, and studies that did not collect rectovaginal swabs to test for GBS, and studies where the full text was not available and not written in English were all excluded.

#### Search Strategy

The review protocol was created by three investigators prior to beginning the study. The electronic databases MEDLINE, ClinicalTrials.gov, PROSPERO, and the Cochrane databse, Wild Card, and Central Register of Controlled Trials were searched from July 2015 of each database until July 2025. The following search criteria were used (“probiotics”[All Fields] OR “probiotical”[All Fields] OR “probiotics”[MeSH Terms] OR “probiotics”[All Fields] OR “probiotic”[All Fields]) AND (“streptococcus agalactiae”[MeSH Terms] OR (“streptococcus”[All Fields] AND “agalactiae”[All Fields]) OR “streptococcus agalactiae”[All Fields] OR “group b streptococcus”[All Fields])”.

#### Data Collection

All articles were independently assessed based on subject, analysis and data extraction based on title, abstract and full text of the articles by two of the investigators, with ties broken by the third. Full-text articles that met inclusion criteria were reviewed in detail, and data were extracted using a standardized template. The following information was collected for each study. Study identification which included first author, year of publication, country of research, study design, study population, sample size, geographical background of participants type, intervention and comparator details [strain(s), and dosage of probiotics used, duration and timing of probiotic administration, comparator group (e.g., placebo, no intervention), and outcome interest]. Differences in the inclusion or exclusion of articles were resolved by vote by the authors. The 2020 PRISMA guidelines were followed for systematic review.

#### Data Items

The primary outcome of interest is the rate of maternal GBS colonization following probiotic administration as assessed by rectal and vaginal swabs. Our secondary outcomes included neonatal GBS infection, preterm birth (< 37 weeks), and perinatal infections (chorioamnionitis and PROM). Data that was closest to delivery or post-intervention and were prioritized. Data collected included participant characteristics: maternal age, gestational age at enrollment, parity, and baseline GBS colonization status, and intervention details. Intervention details include probiotic strain(s), dosage, route of administration, frequency, timing of initiation, and duration of treatment. When critical data was unavailable, attempts were made to contact study authors for clarification.

#### Study Risk of Bias Assessment

We assessed the risk of bias in each included study using the criteria outlined. For randomized controlled trials (RCTs), we used the Cochrane Risk of Bias 2.0 (RoB 2) tool, which evaluates bias across five domains: bias arising from the randomization process, bias due to deviations from intended interventions, bias due to missing outcome data, bias in measurement of the outcome, and bias in selection of the reported result. Each domain was rated as “low risk,” “some concerns,” or “high risk,” and an overall risk of bias judgment was assigned accordingly. Publication bias was evaluated using a funnel plot. Our study was exempt from IRB approval because it collected and integrated publicly available research.

#### Effect measures

For our primary outcome measure of GBS colonization status of positive or negative, GBS neonatal infection, and adverse events we extracted data to compute relative risk ratios with corresponding 95% confidence intervals. When studies reported multiple time points, we prioritized the most recent post-delivery associated data. We ensured that effect measures were converted to match consistency across studies.

#### Reporting Bias assessment

To assess reporting bias, we examined whether the studies were exclusively reporting outcomes of interest or omitted expected outcomes.

#### Certainty Assessment

We incorporated the GRADE assessment tool to assess the certainty of the evidence of primary and secondary outcomes. This approach considered five domains Risk of bias, Inconsistency, Indirectness, Imprecision, and Publication bias. For each outcome, the overall certainty of evidence was rated as high, moderate, low, or very low. Randomized controlled trials started at high certainty and were downgraded based on concerns in any of the five domains. Non-randomized studies started at low certainty but could be upgraded if they demonstrated a large effect size, dose-response relationship, or if all plausible confounding would reduce the observed effect.

Certainty assessments were performed independently by two reviewers, with disagreements resolved through discussion or consultation with a third reviewer. We summarized the findings and GRADE ratings in a Summary of Findings table.

#### Meta-Analysis

A meta-analysis of our primary outcome measure (the rate of maternal GBS colonization following probiotic administration) by using a web-based platform (metaanalysis.com). We calculated Hazard Ratios using an Inverse Variance weighting method on a Random-effects model.

## RESULTS

### Study Selection

The initial search yielded 403 studies, and after removal of duplicates, there were 309 unique studies. 277 of those were excluded, yielding 32 articles for full text retrieval. Two of the authors, DR and AB voted whether studies met inclusion criteria, with one author, KR, breaking ties. Full text articles were reviewed by two reviewers, DR and AB, yielding a total of 14 articles that were unanimously decided to be included. The article selection process is illustrated in Figure 1.

**Figure 1.**
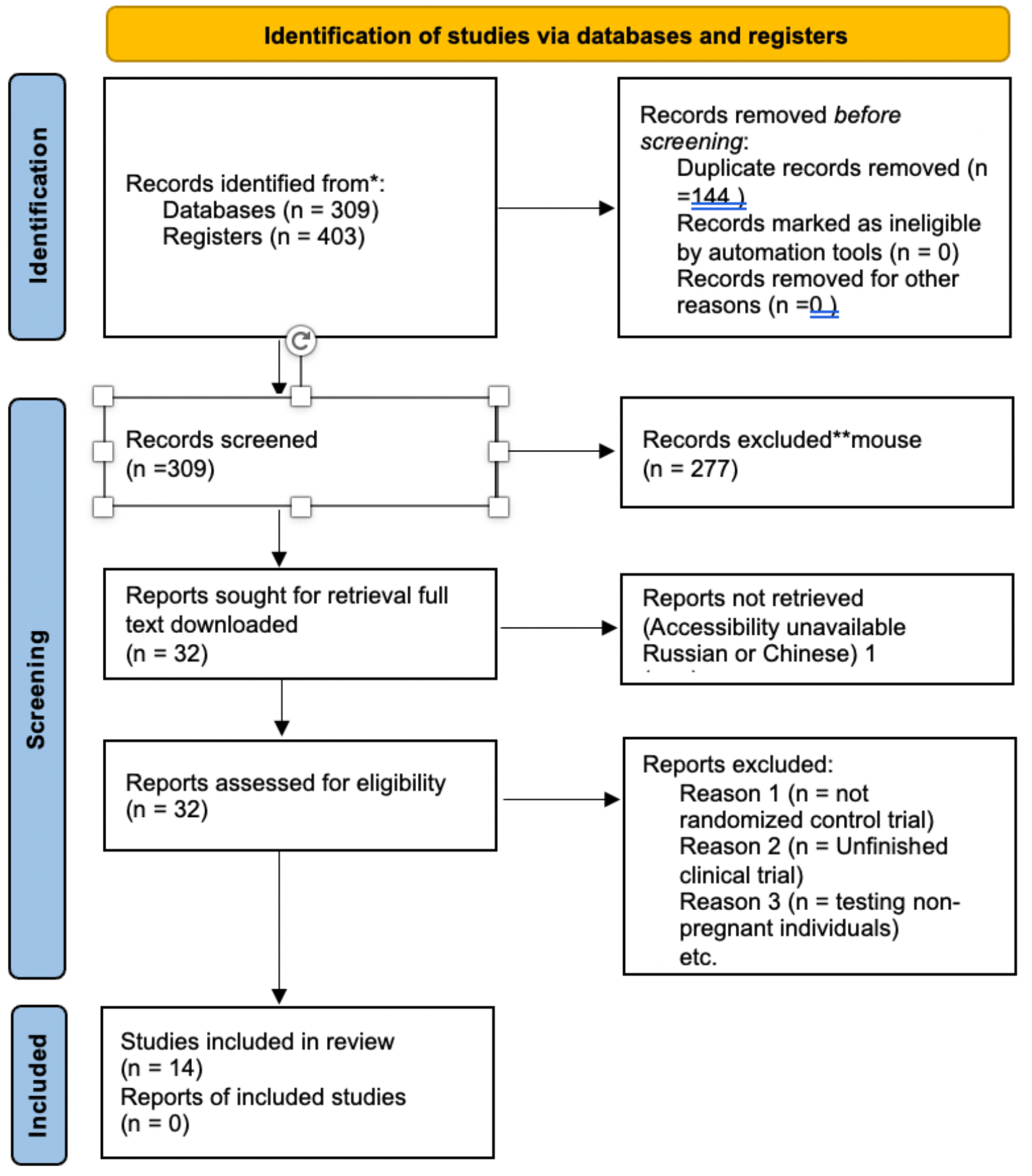
PRISMA Flow Diagram. PRISMA: Preferred Reporting Items for Systematic Reviews and Meta-Analyses.

The 14 studies that met our criteria are summarized in Tables 1, 2 and 3. Table 1 includes data on the study goal, sample size, maternal age and gestational age. Table 2 contains information on GBS status, the probiotic and its administration, and primary outcomes. Table 3 includes information on premature delivery and secondary outcomes. There were a variety of different probiotic formulations used. Interestingly, three studies used the same formulation, Florajen3, that were composed of *Lactobacillus acidophilus, Bifidobacterium lactis* and *B. longum*, and an additional study used Florajen digestion, which contained those three strains plus *B. animalis*.

**Table 1.** Characteristics of Studies Evaluating Oral Probiotic Use for Preventing GBS Colonization in Pregnancy.

| Author, Country, Year | Study Goal/Objective | Sample Size (N) | Maternal Age & Gestational Age (GA) | Administration Route |
| --- | --- | --- | --- | --- |
| Galvez, Spain, 2024[10] | Evaluate percentage of participants with vaginal/rectal GBS colonization at end of intervention (35 wk GA). | 20 | 18–45 y; GA <13 wk | Oral |
| Hanson, USA, 2014 [8] | Examine effect of oral probiotic on GBS colonization and demonstrate feasibility of a larger RCT. | 20 | >18 y; GA 28 ± 2 wk | Oral |
| Hanson, USA, 2023 [9] | Evaluate efficacy of a probiotic to reduce standard-of-care antenatal GBS colonization. | 109 | >18 y; GA 28 ± 2 wk | Oral |
| Ho, Taiwan, 2015 [11] | Examine effect of *L. rhamnosus* GR-1 and *L. reuteri* RC-14 on GBS-positive women becoming GBS negative. | 110 | >18 y; GA 35 ± 2 wk | Oral |
| Kasai, Japan, 2023 [12] | Pilot project to explore causal relationships between oral probiotics and vaginal GBS colonization rates. | 34 | >18 y; GA 35 ± 2 wk | Oral |
| Malloy, USA, 2024 [7] | Compare vaginal/rectal microbiomes before/after probiotic intervention with focus on racial disparities. | 33 | >18 y; GA 28 ± 2 wk | Oral |
| Martin, Spain, 2019 [13] | Evaluate recto-vaginal GBS screening and intrapartum antibiotic prophylaxis (IAP) alternatives. | 54* | 25–35 y; GA not spec. | Oral |
| Menichini, Italy, 2025 [14] | Evaluate feasibility and effects of probiotics starting in 3rd trimester on rectovaginal GBS colonization. | 267 | >18 y; GA 30 wk | Oral |
| Olsen,<br>Australia,<br>2017 [15] | Pilot project to explore causal relationships between oral probiotic use and vaginal GBS rates. | 34 | >18 y; GA<br>36 wk | Oral |
| Di Pierro,<br>Italy,<br>2016 [16] | Assess probiotic use to prevent streptococcal infection while preserving gut microbiota composition. | 406 | >18 y; GA<br>not spec. | Oral |
| Farr,<br>Austria,<br>2020 [17] | Evaluate potential of oral probiotics to eradicate vaginal GBS colonization during 3rd trimester. | 215 | >18 y; GA<br>33 wk | Oral |
| Lai,<br>Taiwan,<br>2021 [18] | Examine effect of *Clostridium butyricum* (Miyarisan BM ) since GA 32+0 on preventing GBS. | 1602 | >18 y; GA<br>32 wk | Oral |
| Sharpe,<br>Canada,<br>2021 [19] | Assess probiotic supplements as a possible way to reduce GBS colonization and IAP side effects. | 139 | 18–45 y;<br>GA<br><25 wk | Oral |
| Nardini,<br>USA,<br>2025 [6] | Determine feasibility of midwife-led RCT of probiotics to reduce GBS colonization by time of birth. | 75 | >18 y; GA<br>33 wk | Oral |

Abbreviations: GA, gestational age; GBS, Group B Streptococcus; IAP, intrapartum antibiotic prophylaxis; RCT, randomized controlled trial.
\*Total sample includes 30 non-pregnant and 24 pregnant patients.

**Table 2.** Probiotic administration and Outcoms on GBS Colonization.

| Author, Country, Year | Initial GBS Status & Timing | Probiotic Strain & Administration | Primary Outcome: Treatment Group [n/N (%)] | Primary Outcome: Control Group [n/N (%)] |
| --- | --- | --- | --- | --- |
| Gálvez, Spain, 2024 | Positive (13 wks) | *Ligalactobacillus. salivarius* (Wks 21–23) | 12/19 (63.2%) Tested negative at 35 wks | 6/22 (27.0%) Tested negative |
| Hanson, USA, 2014 | Unknown (Start) | Florajen3 (*L. acidophilus*, *B. lactis*, *B. longum*) (6 wks from Wk 28) | 8/10 (80.0%) Tested negative (35-37 wks) | 8/10 (80.0%) Tested negative (No placebo) |
| Hanson, USA, 2023 | Unknown (Start) | Florajen3 (6 wks from Wk 28) | 33/39 (84.6%) Tested negative | 35/44 (80.0%) Tested negative (No placebo) |
| Ho, Taiwan, 2015 | Positive (35-37 wks) | *Lactobacillus rhamnosus* GR-1 and *L. reuteri* RC-14(until delivery) | 21/49 (42.9%) Tested negative* | 9/50 (18.0%) Tested negative (Placebo) |
| Kasai, Japan, 2023 | Positive (35-37 wks) | *L. reuteri* (from week 35-37 to a month after delivery) | 22/29 (75.6%) Tested negative at delivery | 25/33 (75.5%) Tested negative (No placebo) |
| Malloy, USA, 2024 | Positive (35-37 wks) | Florajen3 (from Wk 25) | 31/39 (82.0%) Tested negative | 25/33 (75.5%) Tested negative (No placebo) |
| Martín, Spain, 2019 | Positive (Wk 25) | *L. salivarius* (Wks 26–38) | 17/25 (68.0%) Tested negative at 38 wks | 0/14 (0.0%) Tested negative (Placebo) |
| Menichini, Italy, 2025 | Negative/No History | Respecta (*L. acidophilus* and *L. rhamnosus* strains) (Wks 30–37) | 108/133 (81.2%) Tested negative (Rectovaginal) | 113/134 (84.3%) Tested negative (Placebo) |
| Olsen, Australia, 2017 | Positive (Wk 36) | *L. rhamnosus* (until delivery) | 4/19 (21.0%) Tested negative | 3/13 (23.0%) Tested negative (No placebo) |
| Di Pierro, Italy, 2016 | Symptomatic (Vaginitis/Colitis) | *Enterococcus faecium*, *Bifidobacterium animalis*, *L. casei* and *L. lactis* species (weeks 30-40 daily) | 100/127 (78.9%) Tested negative* | 203/279 (72.8%) Tested negative (No placebo) |
| Farr, Austria, 2020 | Positive (Wks 33–37) | Oral Probiotic (Randomized) | 12/33 (36.3%) Tested negative | 6/27 (22.2%) Tested negative (Placebo) |
| Lai, Taiwan, 2021 | Unknown (Start) | *C. butyricum* (4 wks from Wk 32) | 264/299 (88.6%) Tested negative* | 1069/1227 (83.7%) Tested negative (No placebo) |
| Sharpe, Canada, 2021 | < 25 weeks gestation | *L. rhamnosus* (12 weeks) | 48/57 (84.2%) Tested negative | 44/56 (78.6%) Tested negative (Placebo) |
| Nardini, USA, 2025 | Positive (3rd Trimester) | Florajen digestion (*L. acidophilus*, *B. lactis*, *B. animalis*, *B. longum*)(14 days) | 18/25 (72.0%) Tested negative | No difference in perinatal outcomes noted |

**Notes:** n/N indicates number of negative results over total participants. \* Indicates results noted as statistically significant by the authors. Wk(s), Weeks of gestation.

**Table 3.** Secondary Outcomes and Safety Data Including Preterm Delivery and Postpartum Complications.

| Author, Country, Year | Preterm/Premature Delivery Outcomes | Additional Secondary Outcomes |
| --- | --- | --- |
| Gálvez, Spain, 2024 | 2 participants (1 per group); no significant difference | Lower rate of postpartum fever days in probiotic group |
| Hanson, USA, 2014 | Not reported | None reported |
| Hanson, USA, 2023 | 1 case (not tested for GBS; lost to follow-up) | None reported |
| Ho, Taiwan, 2015 | Not reported | 10 newborns admitted to NICU (due to <4h antibiotic prophylaxis) |
| Kasai, Japan, 2023 | Not reported | 15/24 tested GBS positive 1 month postpartum (not significant) |
| Malloy, USA, 2024 | Not reported | None reported |
| Martín, Spain, 2019 | Not reported | None reported |
| Menichini, Italy, 2025 | Not reported | None reported |
| Olsen, Australia, 2017 | Not reported | None reported |
| Di Pierro, Italy, 2016 | Not reported | None reported |
| Farr, Austria, 2020 | No statistically significant difference between groups | None reported |
| Lai, Taiwan, 2021 | Not reported | None reported |
| Sharpe, Canada, 2021 | Not reported | None reported |
| Nardini, USA, 2025 | Not reported | None reported |

Abbreviations: GBS, Group B Streptococcus; NICU, neonatal intensive care unit.

The 14 studies that met our inclusion criteria showed a wide range of efficacy at preventing GBS colonization (Table 2). We performed a meta-analysis on these studies in order to determine overall effectiveness of probiotics at preventing GBS across all the studies (Figure 2). Based on the analysis performed using random effects model with inverse variance method to compare the hazard rate (HR), there is a statistical difference, the summarized hazard ratio (HR) is 0.49 with a 95% confidence interval of 0.31 - 0.78 (Figure 2). The test for overall effect shows a significance at p<0.05. This suggests that probiotics have a significant effect in reducing GBS colonization. A significant heterogeneity was detected (p<0.01), suggesting inconsistent effects in magnitude and/or direction. The I^2^ value indicates that 69% of the variability among studies arises from heterogeneity rather than random chance. A risk of bias assessment is presented in Figure 3, and there was low risk of bias due to randomization, missing outcome data, measurement of the outcome or other factors.

**Figure 2.**
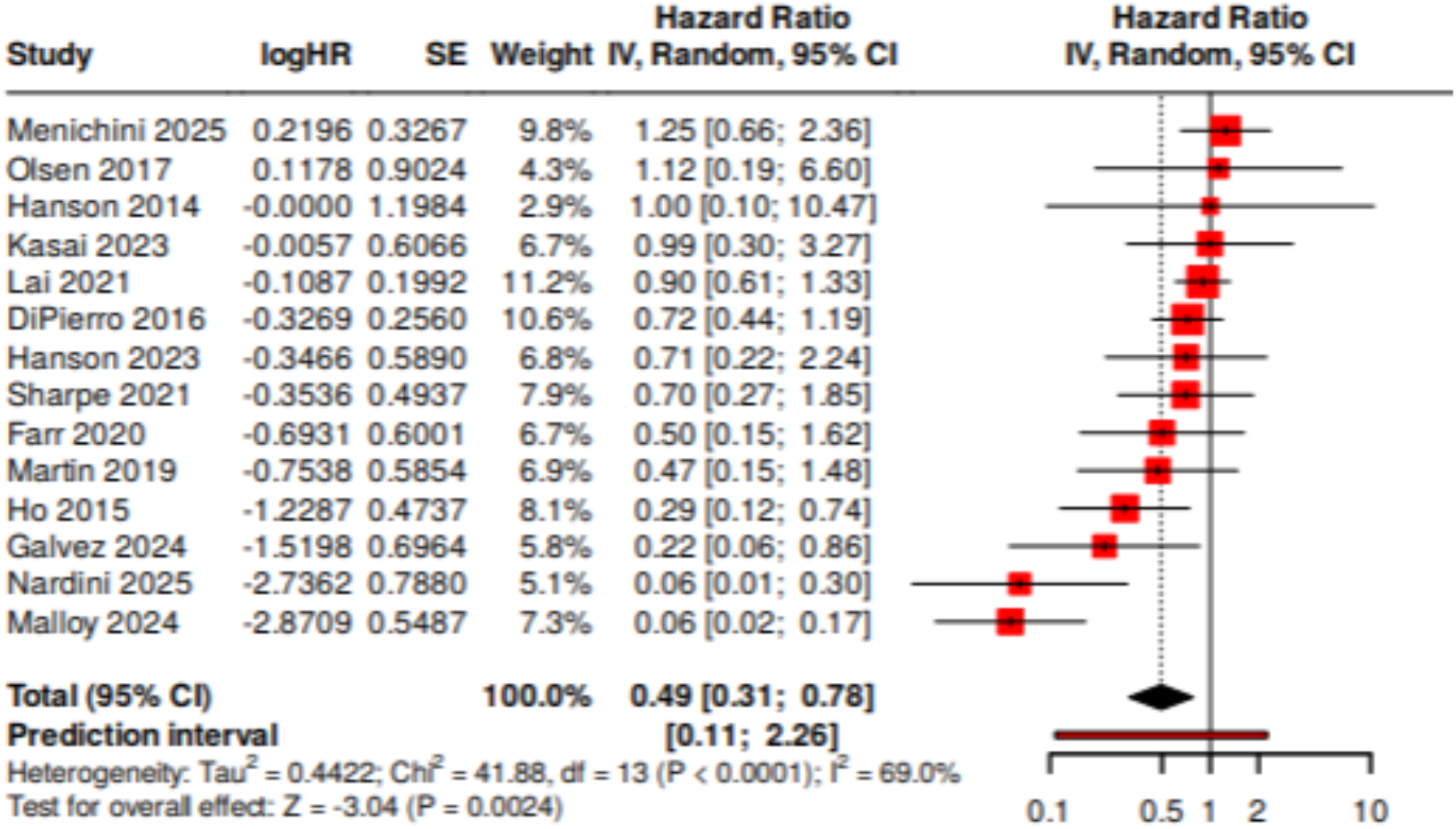
Meta-analysis of effectiveness of probiotics in preventing GBS colonization. Hazard ratios were calculated using an inverse variance weighting method on a random-effects model.

**Figure 3.**
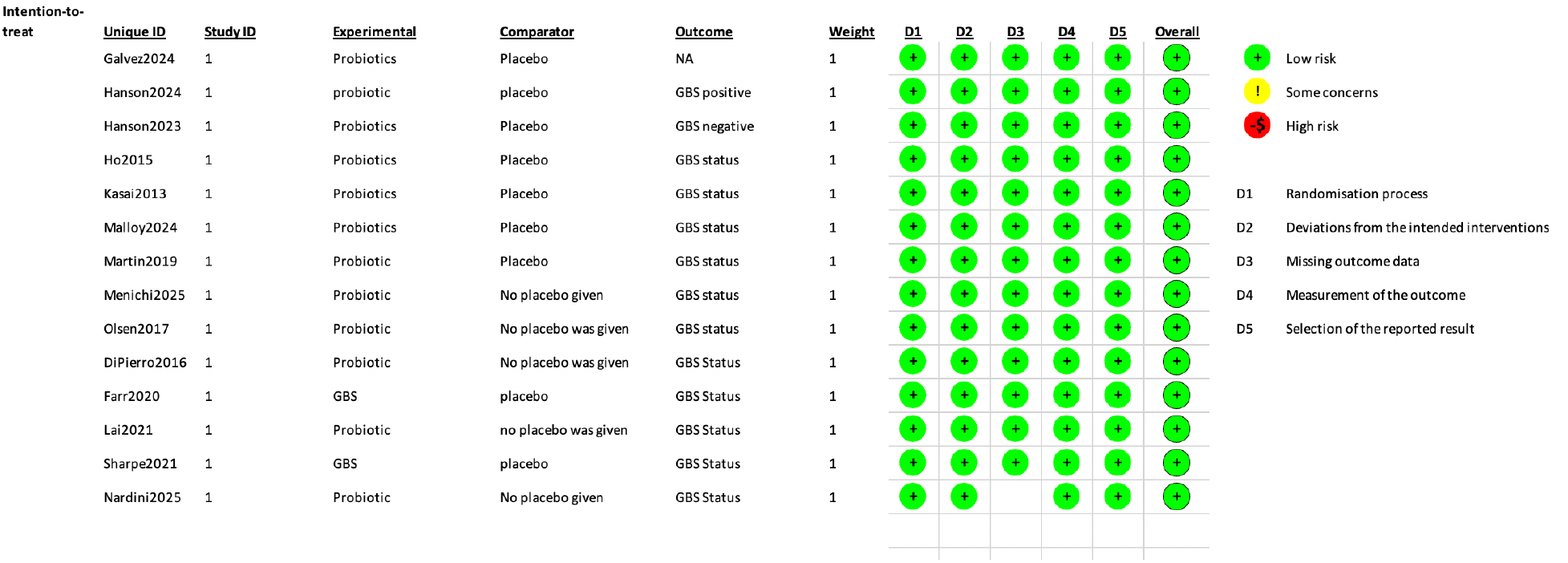
Risk of bias assessment. Each study was assessed for bias due to randomization process, bias in deviations from intended interventions, bias in missing outcome data, bias in measurement of outcomes, and bias in selection of the reported result using the Cochrane Risk of Bias 2.0 tool.

Individually, four of the studies showed dramatic effectiveness of probiotic use in preventing Group B Streptococcus colonization. The other 10 studies showed a range of effectiveness, from partially effective in preventing GBS colonization to no effectiveness. While the meta-analysis showed a significant effect overall, the variance of individual studies suggests that study design may play a role in whether probiotics are effective or not. The two studies that were most effective used Florajen3 as the probiotic [6,7]. However, Hanson 2014 and Hanson 2023 also used Florajen, and their results were inconclusive [8,9]. Most of the studies used similar probiotic bacterial strains, primarily *Lactobacillus* and *Bifidobacterium* strains, but there were no consistent effects of probiotic formulation.

## DISCUSSION

We performed a systematic review and meta-analysis of randomized controlled trials testing whether probiotics prevented GBS colonization in pregnant people. 14 studies met our criteria, and the results showed a variety of effectiveness. There were studies showed that probiotics were dramatically effective in preventing GBS colonization and some showed only partial or very little effectiveness (Figure 2). We performed a meta-analysis, grouping all the included data to test whether probiotics prevented GBS colonization. We found that there was a statistically significant effect in preventing GBS colonization in pregnant individuals across all the studies.

While this systematic review and meta-analysis shows that probiotics are effective in preventing GBS colonization, there is great variability across studies. There were several studies that showed significant probiotic effects [6,7,10,11] but several had no effect at all [6,13,15].

Although the meta-analysis revealed a statistically significant overall effect, the variability among individual study results highlights the importance of study design, population characteristics, and probiotic formulation in determining efficacy. Notably, the two studies that reported the most effective outcomes utilized Florajen3 and Florajen digestion, which were similar multi-strain probiotics. Florajen 3 was composed of *Lactobacillus acidophilus, Bifidobacterium lactis*, and *Bifidobacterium longum*, while Florajen digestion contained strains from those three species along with *Bifidobacterium animalis* [6,7]. Interestingly, two other studies using Florajen 3 showed minimal effects in reducing GBS colonization [8,9]. These results suggest that consistency in strain selection, dosage, and duration of administration could be critical factors influencing success. Taken together, these findings underscore both the promise of probiotics as a preventive strategy against GBS colonization and the need for further standardized, large-scale clinical trials to clarify their role in obstetric care.

Once a consistent probiotic regimen that is effective at clearing GBS is determined, it could be a great benefit for antibiotic stewardship. Current guidelines suggest testing pregnant individuals for GBS, and if there is a positive test, treat with intrapartum intravenous penicillin. With an improved regimen, pregnant individuals who test positive could be treated with oral probiotics, and a follow up test performed to test effectiveness. In the United States alone, there are more than 3 million live births each year. If 25% of those individuals receive antibiotics, the use of probiotics could reduce antibiotic usage by a significant amount. Greater antibiotic stewardship would reduce the likelihood of the development of antibiotic resistance and would be less disruptive to the microbiome of the individuals involved.

## Data Availability

All data produced in the present work are contained in the manuscript

## ACKNOWLEDGMENTS

Gemini was used for the sole purpose of formatting tables. It was not used for data analysis or writing of the manuscript.

